# Regression-Based Balance Discordance Refines Prediction of Prospective Falls After Subacute Stroke: A Secondary Analysis of the LEAPS Trial

**DOI:** 10.64898/2026.11.06.26367447

**Authors:** Andrew Hooyman, Daniel S. Peterson, Jason K. Longhurst, Franziska Albrecht

**Affiliations:** Department of Physical Therapy, Chapman University, Irvine, CA, USA; College of Health Solutions, Arizona State University, Phoenix, AZ, USA; Department of Physical Therapy and Athletic Training, Saint Louis University, Saint Louis, MO, USA; Division of Physiotherapy, Department of Neurobiology, Care Sciences and Society, Karolinska Institutet, Stockholm, Sweden; Women’s Health and Allied Health Professional Theme, Karolinska University Hospital, Stockholm, Sweden

## Abstract

**Objective:** To determine whether continuous, regression-based measures of balance discordance refine prediction of prospective, recurrent falls after subacute stroke relative to a previously published threshold-based classification in the same cohort, and to characterize how a purely data-driven, dimensionality-reduction-based composite expresses the same underlying measures.

**Design:** Secondary, observational analysis of a randomized controlled trial. Recurrent falls were modeled using Andersen-Gill Cox proportional hazards regression with robust standard errors clustered by participant.

**Setting:** Five-site, National Institutes of Health/National Institute of Neurological Disorders and Stroke funded multicenter trial (Locomotor Experience Applied Post-Stroke, LEAPS).

**Participants:** 384 adults with subacute stroke (of 408 randomized in the parent trial) with valid baseline balance confidence, walking, and balance scores and at least one recorded follow-up interval (2,357 person-intervals, 179 fall events).

**Main Outcome Measures:** Prospective, recurrent falls over 12 months, ascertained via self-report falls diary. Balance discordance was computed as the residual from regressing balance confidence on objective walking distance, Berg Balance Scale performance, or both jointly (multivariable discordance); threshold-based classification used established cut-points on the same measures and principal component analysis served as secondary comparators.

**Results:** Single-variable discordance was associated with lower fall hazard (hazard ratio [HR] = 0.79, 95% CI 0.63-0.99, p = .037), and multivariable discordance was associated with higher fall hazard (HR = 1.27, 95% CI 1.03-1.58, p = .026). None of three threshold-based classification indicators reached significance. Multivariable discordance and a three-component principal-component composite showed comparable model fit (concordance 0.616 vs. 0.618) and were substantially correlated (R^2^ = .49).

**Conclusions:** Continuous, regression-based balance discordance predicted prospective, recurrent falls where a previously published threshold-based classification did not. The divergent direction of single- and multivariable findings suggests overconfidence relative to combined objective function, rather than confidence broadly, may be the more specific marker of fall risk.

## Introduction

Falls are among the most common and consequential complications of stroke recovery, with recurrent falls documented in roughly a quarter to a third of survivors within the first year and continuing well beyond the acute phase.^1,2^ Identifying which survivors are at greatest risk, and doing so early enough to inform rehabilitation planning, remains a persistent clinical challenge, since objective impairment alone does not fully account for who ultimately falls.^3,4^ Balance confidence, measured independently of objective balance or gait performance, has been linked to functional recovery, participation, and mobility outcomes after stroke,^5,6^ yet its relationship to falls specifically has been inconsistent: some cohorts find balance confidence predictive of future falls, while others find no independent association once objective function is accounted for.^4^ This inconsistency suggests that perceived confidence and objective performance, considered separately, each capture only part of the picture relevant to fall risk.

An alternative approach considers not confidence or performance in isolation, but the mismatch between them. Using a threshold-based approach, early work in community-dwelling older adults classified individuals by whether their perceived and physiological fall risk each fell above or below established cut-points. This approach separates individuals into 4 quadrants, 2 aligned or “concordant” (high confidence/performance, low confidence/performance) and 2 misaligned or “discordant” (overconfident, underconfident). Indeed, discordance of balance confidence and balance performance, rather than either measure alone, related meaningfully to psychological and behavioral correlates of falling.^7^ In stroke specifically, a similar threshold-based approach was applied to the Locomotor Experience Applied Post-Stroke (LEAPS) trial cohort,^8,9^ classifying participants by whether their balance confidence (Activities-Specific Balance Confidence Scale)^10^ and balance performance (Berg Balance Scale)^11^ fell above or below established cut-points.^12^ That analysis found misalignment of balance confidence relative to high balance ability was associated with subsequent gait speed, but did not find a clear association with fall incidence.

Our group has since developed a continuous, regression-based approach to quantifying discordance, in which balance confidence is regressed on objective balance or mobility performance and the resulting residual, rather than a categorical cut-point, serves as the discordance score. In Parkinson’s disease, this approach has predicted retrospective faller status via logistic regression across multiple independent cohorts, with greater discordance (confidence exceeding objectively predicted confidence) associated with lower odds of having previously fallen.^13,14^ Follow-up work further showed that discordance also relates to non-fall outcomes such as quality of life and cognition, and to pre–post change with intervention.^15–17^

These continuous quantifications of misalignment are meaningfully related to functional outcomes. However, the single-variable formulation is unidimensional with respect to participant function. Function can be expressed in several ways (i.e. gait speed, walking capacity, balance, etc.) and each can be independently linked to balance confidence, so regressing balance confidence on any one measure yields a narrow expression of the gap between perceived and actual ability. The brain-age-gap literature^18^ offers a methodology for a multivariable alternative, in which multiple objective measures are fit jointly to a single self-report, producing a more robust and potentially more representative index of misalignment; this approach has not yet been applied to balance.

The primary purpose of the present study is to determine whether continuous, regression-based approaches to quantifying balance discordance refine the prediction of prospective falls, and offers insight into their possible source, beyond the previously published threshold-based classification provided in this same cohort.^12^ In the LEAPS trial cohort we modeled prospective falls with recurrent-event survival analysis with different formulations of discordance as the predictive exposure to address four aims: (1) determine whether regression-based discordance improves prediction of prospective falls relative to the previously published threshold-based classification in this same cohort; (2) determine whether single-variable balance discordance predicts the hazard of prospective falls; (3) determine whether multivariable balance discordance, incorporating two objective measures, does so as well; and (4) establish how multivariable discordance relates to a purely data-driven, dimensionality-reduction-based composite (principal component analysis), clarifying how each approach partitions shared and distinct variance across measures.

## Methods

### Design and Participants

This study is a secondary, observational analysis of data from the LEAPS trial, a five-site, three-arm, single-blind, National Institutes of Health/National Institute of Neurological Disorders and Stroke funded phase III randomized controlled trial (Duncan et al., 2007, 2011). Participants were adults aged 18 years or older, 5 to 30 days post-stroke at enrollment, who were randomized at 2 months post-stroke to early locomotor training, late locomotor training, or a home exercise program. Assessments occurred at screening and at 2-, 6-, and 12-months post-stroke. In the present analysis, randomized treatment group was retained solely as an adjustment covariate, consistent with this study’s observational, non-causal aim.

Of the 408 participants randomized in the parent trial (Figure 1), 401 had a valid 2-month Activities-specific Balance Confidence score and valid fall status and were retained to compute discordance and comparator scores. After merging with the falls diary and restricting to person-intervals up through each participant’s first recorded fall event, the final analytic sample for all survival models comprised 384 participants, contributing 2,357 person-intervals and 179 fall events (approximately 9.4 events per model parameter).

**Figure 1.**
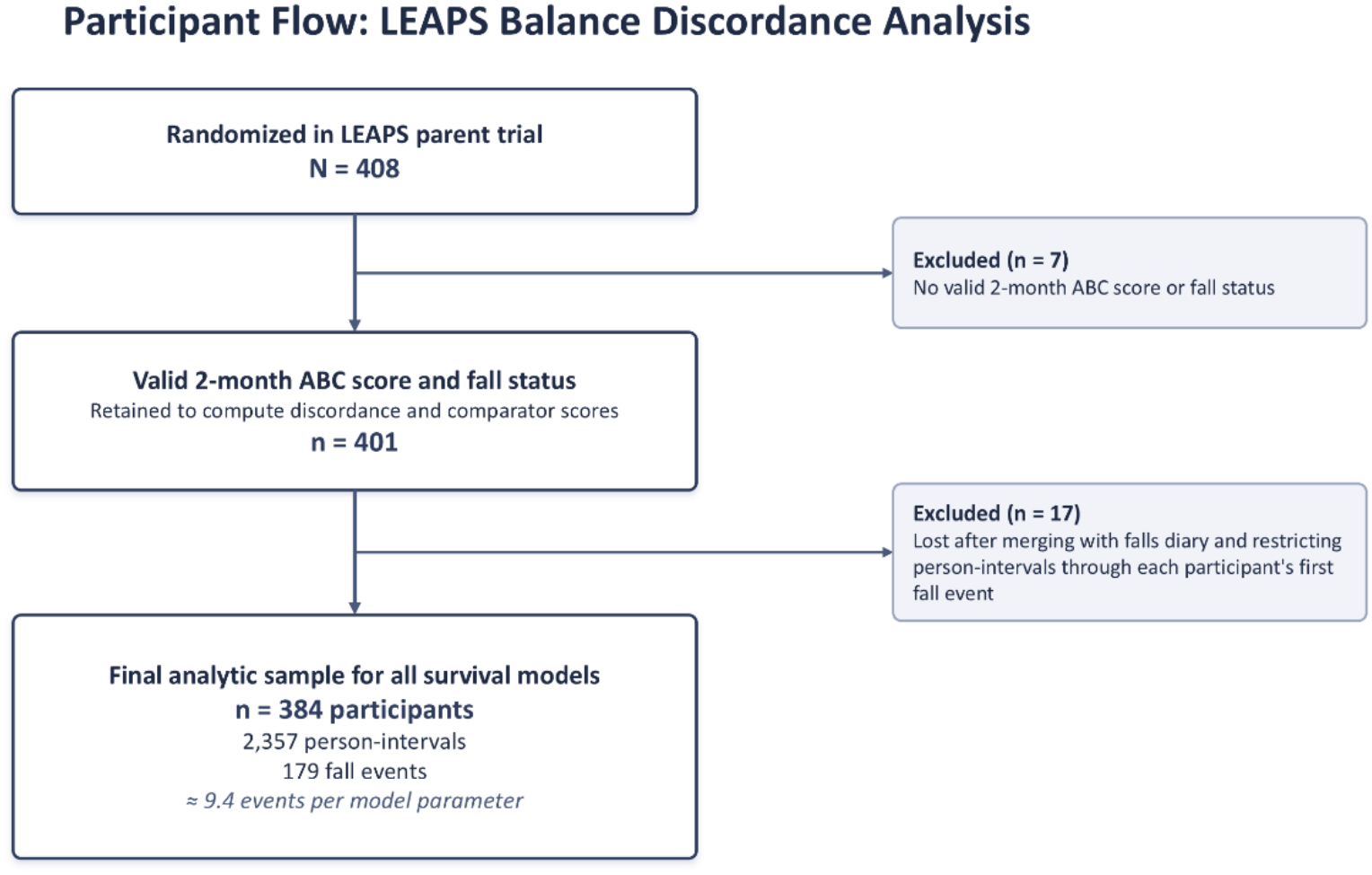
Participant flow for the balance discordance analysis of the LEAPS trial. Of the 408 participants randomized in the parent trial, 401 had a valid 2-month Activities-specific Balance Confidence (ABC) score and valid fall status and were retained to compute discordance and comparator scores. After merging with the falls diary and restricting person-intervals to those occurring up through each participant’s first recorded fall event, the final analytic sample for all survival models comprised 384 participants, who contributed 2,357 person-intervals and 179 fall events (approximately 9.4 events per model parameter). *Abbreviation:* ABC, Activities-specific Balance Confidence.

### Measures

Balance confidence was assessed with the Activities-specific Balance Confidence Scale.^10^ Objective walk capacity and balance performance were assessed with the 6-Minute Walk Test distance^19^ and the Berg Balance Scale.^11^ All three measures were collected at the 2-month baseline assessment.

Single-variable balance discordance was computed separately for each objective measure. Balance confidence was regressed on one objective measure, walking capacity or Berg Balance Scale performance, and the residual served as the discordance score: the difference between observed confidence and the confidence predicted from that measure. A multivariable balance discordance score was computed from a joint regression of balance confidence on both objective measures, followed by a bias-correction step adapted from the age-level correction approach used in brain-age-gap research (Zhang et al., 2023),^18^ in which the joint model’s predicted confidence values were regressed on observed confidence to correct for regression-to-the-mean compression in the prediction before computing the final residual. Positive discordance values indicate confidence exceeding what objective performance would predict (overconfidence); negative values indicate confidence lower than what objective performance would predict (underconfidence).

As a methodological comparator, principal component analysis was conducted on the centered and scaled 2-month balance confidence, walking distance, and Berg Balance Scale scores, yielding a symmetric composite in which no single measure is privileged as an outcome.

As a second comparator, threshold-based binary classification variables were derived using established cut-points: balance confidence of 66 or below and Berg Balance Scale score of 42 or below, following Liphart and colleagues (2016),^12^ and walking distance of 288 m or below, following Fulk and colleagues (2017)^20^, who derived this cut-point (distinguishing limited from unlimited community ambulators) from a pooled stroke sample that included the LEAPS cohort.

Prospective falls were ascertained via a self-report falls diary. Participants received a definition of a fall, 30-day calendars, and were instructed to mail a postcard when a fall occurred; monthly phone contacts also captured fall occurrence, and a structured phone call characterized each reported fall.

### Statistical Analysis

Falls were modeled as recurrent events using the Andersen-Gill extension of the Cox proportional hazards model, with a counting-process (start, stop, event) data structure and robust standard errors clustered by participant to account for within-person correlation across repeated fall intervals. All models adjusted for age, randomized treatment group, sex, race, ethnicity, stroke location, stroke type, days from stroke onset to randomization, cognitive status (Mini-Mental State Examination), depressive symptoms (Patient Health Questionnaire-9), and lower-extremity motor impairment (Fugl-Meyer).

Four models were fit. **Model 1 (primary)** entered three terms jointly: the more sensitive single-variable discordance score in this cohort, the multivariable discordance score, and the three threshold-based classification indicators. This tested directly whether continuous discordance predicts fall hazard beyond threshold-based classification, while estimating each term’s independent association. **Model 2** repeated Model 1 but substituted the alternative single-variable discordance score, as a consistency check. **Models 3 and 4** addressed a secondary, methodological comparison between multivariable discordance and PCA, clarifying how each expresses the shared and distinct variance among the measures rather than making an independent claim about fall prediction: Model 3 entered both discordance terms (no threshold or PCA terms); Model 4 entered all three principal components (no discordance or threshold terms). Discordance and principal component terms were never entered into the same model, and the two single-variable discordance candidates were never entered together, since each pairing would introduce substantial collinearity given their shared derivation from balance confidence scores. Models were compared on Akaike Information Criterion (AIC) and concordance where relevant. All discordance and principal component terms were standardized (mean-centered, scaled to unit variance) prior to entry, so that hazard ratios reflect the change in hazard per 1-SD increase. Variance inflation factors were examined for multicollinearity, and the proportional hazards assumption was assessed using scaled Schoenfeld residuals. All tests used a conventional two-sided alpha of .05.

This analysis was preregistered prior to any outcome-linked analysis (Open Science Framework, https://osf.io/j3cqu/overview). A supplementary statistical analysis document provides the complete analytic code and maps each model and results directly to the corresponding preregistered research question and hypothesis.

## Results

The analytic sample comprised 384 participants (2,357 person-intervals, 179 fall events); its derivation from the 408 randomized is shown in Figure 1. Participant characteristics are presented in Table 1.

**Table 1.** Baseline Characteristics of the Analytic Sample (N = 384)

| **Characteristic** | **N = 384** |
|---|---|
| Age, years | 62.1 (12.8) |
| **Sex** |  |
| Male | 209 (54%) |
| Female | 175 (46%) |
| **Race** |  |
| American Indian/Alaska Native | 5 (1.3%) |
| Asian | 51 (13%) |
| Black/African American | 85 (22%) |
| White | 220 (57%) |
| Native Hawaiian/Pacific Islander | 19 (4.9%) |
| More than one race | 3 (0.8%) |
| Unknown | 1 (0.3%) |
| **Ethnicity** |  |
| Hispanic/Latino | 59 (15%) |
| Not Hispanic/Latino | 325 (85%) |
| **Randomized group** |  |
| Early Training | 129 (34%) |
| Late Training | 133 (35%) |
| Home Exercise | 122 (32%) |
| Days from stroke to randomization | 63.7 (8.2) |
| **Stroke location** |  |
| Right hemisphere | 182 (47%) |
| Left hemisphere | 138 (36%) |
| Brainstem | 58 (15%) |
| Bilateral | 6 (1.6%) |
| **Stroke type** |  |
| Ischemic | 307 (80%) |
| Hemorrhagic | 72 (19%) |
| Uncertain/unclassified | 5 (1.3%) |
| MMSE, baseline | 26.0 (3.5) |
| Missing | *35* |
| PHQ-9 depression, baseline | 4.6 (5.2) |
| Lower-extremity Fugl-Meyer, baseline | 21.1 (6.4) |
| Missing | *1* |
| ABC scale, baseline (%) | 45.1 (23.8) |
| 6-Minute Walk Test, baseline (m) | 126.5 (78.2) |
| Berg Balance Scale, baseline | 35.8 (14.0) |
| Below ABC cut-point (66 or less) | 306 (80%) |
| Below 6MWT cut-point (288 m or less) | 375 (98%) |
| Below Berg cut-point (42 or less) | 230 (60%) |
| **Liphart (2016) 4-group discordance classification** |  |
| Group 1: Concordant (low ABC/low BBS) | 201 (52%) |
| Group 2: Discordant (high ABC/low BBS) | 29 (7.6%) |
| Group 3: Concordant (high ABC/high BBS) | 49 (13%) |
| Group 4: Discordant (low ABC/high BBS) | 105 (27%) |
| Single-variable discordance, 6MWT-based | -0.1 (21.9) |
| Single-variable discordance, Berg-based | 0.0 (21.3) |
| Multivariable discordance, decorrelated | 0.0 (46.3) |
| **Fall during follow-up** |  |
| No fall | 188 (49%) |
| 1+ fall | 196 (51%) |

*Note. Values are mean (SD) or n (%). ABC = Activities-specific Balance Confidence Scale; BBS = Berg Balance Scale; MMSE = Mini-Mental State Examination; PHQ-9 = Patient Health Questionnaire-9; 6MWT = 6-Minute Walk Test.*

### Single-Variable and Multivariable Discordance

In the primary model, which entered the more sensitive single-variable discordance candidate (Berg Balance scale-derived), the multivariable discordance score, and the three threshold-based classification indicators jointly, single-variable discordance was significantly associated with the hazard of prospective falls (hazard ratio [HR] = 0.79, 95% confidence interval [CI] 0.63-0.99, p = .037): greater discordance, reflecting confidence exceeding what objective performance would predict, was associated with lower fall hazard. Multivariable discordance was also significantly associated with fall hazard in this model (HR = 1.27, 95% CI 1.03-1.58, p = .026), in the opposite direction: greater multivariable discordance was associated with higher fall hazard. Model concordance was 0.62, and the robust score test, which accounts for the clustering of repeated fall intervals within participants, was significant (p = .01).

A consistency-check model substituting the alternative single-variable discordance candidate (6MWT derived) showed the same direction of association (HR = 0.81) but did not reach significance (p = .059); the multivariable discordance score remained directionally consistent in this model (HR = 1.21).

### Threshold-Based Classification

None of the three threshold-based classification indicators, entered alongside the continuous discordance scores in the primary model, was independently significant (ABC threshold: HR = 0.74, p = .18; 6MWT threshold: HR = 0.55, p = .068; Berg Balance Scale threshold: HR = 1.42, p = .070), although the latter two showed trend-level associations. By contrast, both continuous discordance terms reached significance in the same model. Visualization of hazard ratios for each discordance measure in Figure 2.

**Figure 2.**
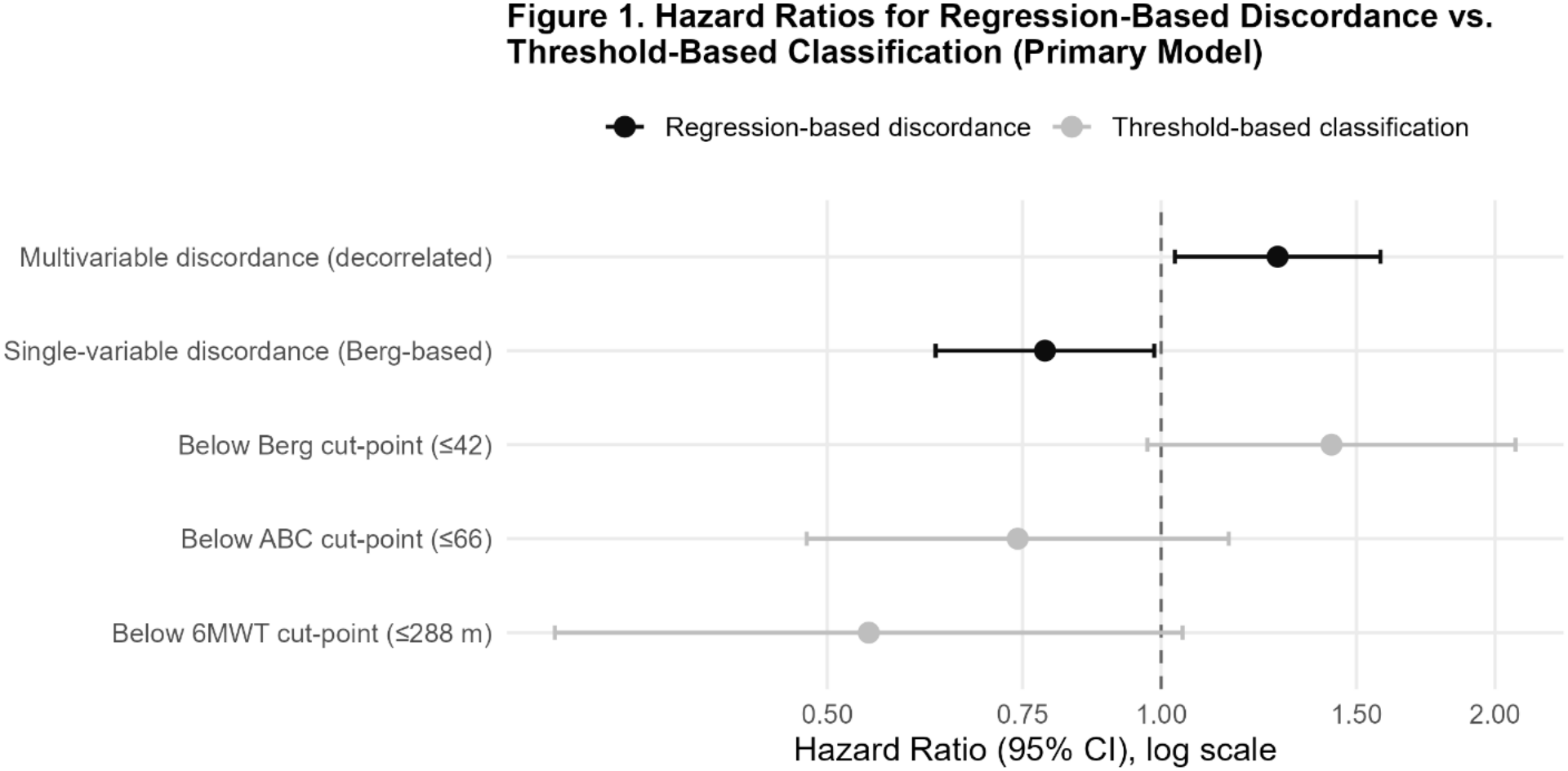
Hazard ratios for regression-based discordance versus threshold-based classification of balance confidence, walking distance, and balance scale performance in predicting prospective, recurrent falls after subacute stroke (primary model). Points represent hazard ratios and horizontal bars represent 95% confidence intervals from a recurrent-event (Andersen-Gill) Cox proportional hazards model adjusted for age, randomized treatment group, sex, race, ethnicity, stroke location, stroke type, days from stroke onset to randomization, cognitive status, depressive symptoms, and lower-extremity motor impairment (N = 384; 2,357 person-intervals; 179 fall events). The vertical dashed line at 1.0 indicates no association with fall hazard. Blue points denote the two continuous, regression-based discordance measures; red points denote the three threshold-based classification indicators built from the same underlying measures. ABC = Activities-specific Balance Confidence Scale; Berg = Berg Balance Scale; 6MWT = 6-Minute Walk Test.

### Multivariable Discordance and the Principal Component Comparator

A model entering both discordance terms without threshold or principal component terms showed a similar pattern to the primary model: single-variable discordance HR = 0.81 (95% CI 0.69-0.96), multivariable discordance HR = 1.36 (95% CI 1.17-1.59), both significant. A parallel model entering all three principal components in place of the discordance terms showed the first principal component associated with lower fall hazard (HR = 0.75) and the second and third components showing no meaningful association (HR = 0.99 and 0.96, respectively). Model fit was nearly identical between the two approaches (discordance model: AIC = 1962.7, concordance = 0.616; principal-component model: AIC = 1964.6, concordance = 0.618). The multivariable discordance score and the first principal component were substantially correlated (R^2^= .49), consistent with both being derived from the same underlying covariance structure among the three input measures rather than capturing independent information. The first principal component explained 68.9% of the total variance, with comparable loadings across the three input measures (balance confidence = 0.49, walking distance = 0.61, Berg Balance Scale = 0.62).

### Model Diagnostics

Variance inflation factors were below 5 in all models, indicating acceptable multicollinearity among covariates and focal predictors. The global proportional hazards assumption was met in all models (p = .21-.25); an individual covariate, stroke type, showed a significant violation of this assumption in two models (p = .007), which is addressed in the Study Limitations section.

## Discussion

This study extended a continuous, regression-based approach to quantifying balance discordance to stroke, and to prospective, recurrent falls, for the first time in any population. Both single-variable and multivariable discordance significantly predicted the hazard of prospective falls in this cohort, with single-variable discordance associated with lower fall hazard and multivariable discordance associated with higher fall hazard. Neither of the threshold-based classification indicators, built from the same underlying measures, reached significance, consistent with a previously published categorical analysis of discordance in this same cohort that similarly found no clear association with fall incidence.^12^ Together, these findings suggest that a continuous representation of the mismatch between perceived and objective function captures fall-relevant information that a categorical cut-point-based representation of the same measures does not.

The divergent direction of the single-variable and multivariable findings warrants explanation. A substantial body of prior work supports balance confidence, considered on its own, as a protective factor: greater confidence has been associated with lower prospective fall risk in community-dwelling older adults independent of age and fall history,^21^ and with better subsequent gait speed in this same population.^22^ This is consistent with the present study’s single-variable finding. However, a separate body of work indicates that confidence specifically exceeding what one’s actual functional ability, rather than confidence in general, is associated with greater fall risk. In a nursing-home sample, residents whose fall-related self-efficacy was dissociated from (exceeded) their measured functional independence fell more often than those whose confidence and function were aligned,^23^ and an interaction between high falls efficacy and poor postural balance was associated with greater fall risk than the same poor balance paired with lower efficacy.^22^ In stroke specifically, a discrepancy between the walking speed a patient chose for a short distance and the speed they could sustain over 6 minutes, an overestimation of endurance capacity, was the strongest predictor of falls in the year after a subacute-stage patient returned home.^24^ Overestimation of functional mobility (Timed Up and Go) performance has been associated with worse objective balance scores in stroke.^25^ Errors in perceiving one’s own postural reach limits have similarly predicted recurrent falls among hospitalized stroke inpatients.^26^ Overestimation of a specific physical ability, not confidence broadly, has similarly been linked to falls in general older-adult samples.^27^

Taken together, this literature suggests a plausible account for the present findings: balance confidence considered relative to a single objective measure may still largely reflect a generally protective, engagement-supporting construct. However, confidence considered relative to the fuller picture of objective function captured by multiple measures together may more precisely isolate the specific miscalibration, overconfidence relative to one’s actual combined balance and mobility capacity, that other work has linked to elevated fall risk. Impaired self-awareness of motor deficits after stroke (anosognosia for hemiplegia) offers one plausible, stroke-specific mechanism by which such miscalibration could arise, though this was not directly assessed in the present study and this connection should be treated as speculative.^28^

A methodological account of the opposing signs is more cautious than a conceptual one. Raw residual scores remain mechanically dependent on the measure regressed out, so the uncorrected single-variable score stays partly confounded with balance confidence, and its protective association (HR = 0.79) may reflect that dependence rather than discordance itself.^29,30^ The bias-corrected multivariable score removes this dependence, isolating confidence unsupported by either domain and yielding the opposing, risk-direction association (HR = 1.27). Whether such correction can reverse an association’s direction, rather than attenuate a confounded one, has not been characterized in the brain-age literature;^31^ we therefore present this divergence as specific to these data.

The comparison between multivariable discordance and the principal component analysis addressed a different kind of question: not which approach predicts falls better, but what each approach represents. The two approaches showed nearly identical model fit and were substantially correlated, consistent with both being derived from the same underlying covariance structure among the same three measures; neither can extract more fall-relevant information than exists in that shared structure. Given this parity in predictive performance, the case for the discordance approach rests on interpretability rather than superior prediction: a discordance score has a fixed, directional clinical meaning, indicating whether a person’s confidence exceeds or falls short of what their objective function predicts, whereas a principal component is a data-driven composite whose meaning depends on inspecting its loadings and is not guaranteed to generalize its exact form to another sample.

### Study Limitations

Several limitations warrant consideration. The proportional hazards assumption was violated for one covariate, stroke type, in two of the four models; while the global test of proportionality was not significant in any model, this individual violation suggests that the effect of stroke type on fall hazard may not be constant over the follow-up window, and results involving stroke type should be interpreted cautiously. The originally published threshold-based classification for this cohort^12^ could not be modeled directly in its original four-group form because one resulting subgroup was too small to support stable estimation and introduced substantial collinearity with the continuous discordance scores. Three separate binary threshold indicators were used instead, and the original four-group classification is reported descriptively only. The mechanistic explanation offered above for the divergent single- and multivariable findings is plausible given prior literature but was not directly tested; this study did not assess awareness of motor deficits or related cognitive constructs. Finally, as a secondary analysis of a completed trial, this study’s covariate set and follow-up window were constrained by the parent trial’s original design.

### Conclusion

Continuous, regression-based measures of balance discordance predicted prospective, recurrent falls in subacute stroke, extending this approach beyond Parkinson’s disease and beyond retrospective fall history for the first time, and doing so where a previously published categorical approach in this same cohort did not. The divergent directions of the single- and multivariable findings point toward overconfidence relative to combined objective function, rather than confidence in general, as the more specific marker of fall risk, and future work should test this distinction directly.

## Data Availability

This study is a secondary analysis of de-identified data from the Locomotor Experience Applied Post-Stroke (LEAPS) trial (ClinicalTrials.gov NCT00243919). The LEAPS trial was approved by the Institutional Review Boards of all participating centers, and all participants provided written informed consent. The de-identified dataset is archived in the National Institute of Neurological Disorders and Stroke (NINDS) data repository and was obtained with the written permission of the parent trial's Principal Investigator for the purposes of this analysis. The present secondary analysis of de-identified data was determined not to constitute human subjects research and was granted exemption by the Chapman University Institutional Review Board.

## Ethics Approval

This study is a secondary analysis of de-identified data from the Locomotor Experience Applied Post-Stroke (LEAPS) trial (ClinicalTrials.gov NCT00243919). The LEAPS trial was approved by the Institutional Review Boards of all participating centers, and all participants provided written informed consent.^9^ The de-identified dataset is archived in the National Institute of Neurological Disorders and Stroke (NINDS) data repository and was obtained with the written permission of the parent trial’s Principal Investigator for the purposes of this analysis. The present secondary analysis of de-identified data was determined not to constitute human subjects research and was granted exemption by the Chapman University Institutional Review Board.

## References

1. Minet, L. R., Peterson, E., von Koch, L. & Ytterberg, C. Occurrence and Predictors of Falls in People With Stroke: Six-Year Prospective Study. Stroke 46, 2688–2690 (2015).

2. Samuelsson, C. M.Hansson, P.-O. & Persson, C. U. Determinants of Recurrent Falls Poststroke: A 1-Year Follow-up of the Fall Study of Gothenburg. Arch. Phys. Med. Rehabil. 101, 1541–1548 (2020).

3. Mansfield, A. et al. Do measures of reactive balance control predict falls in people with stroke returning to the community? Physiotherapy 101, 373–380 (2015).

4. Simpson, L. A., Miller, W. C. & Eng, J. J. Effect of stroke on fall rate, location and predictors: a prospective comparison of older adults with and without stroke. PloS One 6, e19431 (2011).

5. Schmid, A. A. et al. Balance and balance self-efficacy are associated with activity and participation after stroke: a cross-sectional study in people with chronic stroke. Arch. Phys. Med. Rehabil. 93, 1101–1107 (2012).

6. Torkia, C., Best, K. L., Miller, W. C. & Eng, J. J. Balance Confidence: A Predictor of Perceived Physical Function, Perceived Mobility, and Perceived Recovery 1 Year After Inpatient Stroke Rehabilitation. Arch. Phys. Med. Rehabil. 97, 1064–1071 (2016).

7. Delbaere, K., Close, J. C. T., Brodaty, H., Sachdev, P. & Lord, S. R. Determinants of disparities between perceived and physiological risk of falling among elderly people: cohort study. BMJ 341, c4165 (2010).

8. Duncan, P. W. et al. Protocol for the Locomotor Experience Applied Post-stroke (LEAPS) trial: a randomized controlled trial. BMC Neurol. 7, 39 (2007).

9. Duncan, P. W. et al. Body-weight-supported treadmill rehabilitation after stroke. N. Engl. J. Med. 364, 2026–2036 (2011).

10. Powell, L. E. & Myers, A. M. The Activities-specific Balance Confidence (ABC) Scale. J. Gerontol. A. Biol. Sci. Med. Sci. 50A, M28–34 (1995).

11. Berg, K. O., Maki, B. E., Williams, J. I., Holliday, P. J. & Wood-Dauphinee, S. L. Clinical and laboratory measures of postural balance in an elderly population. Arch. Phys. Med. Rehabil. 73, 1073–1080 (1992).

12. Liphart, J. et al. Concordance and discordance between measured and perceived balance and the effect on gait speed and falls following stroke. Clin. Rehabil. 30, 294–302 (2016).

13. Longhurst, J. K. et al. Discordance Between Balance Ability and Perception Is Associated With Falls in Parkinson’s Disease: A Coordinated Analysis. Neurorehabil. Neural Repair 39, 114–125 (2025).

14. Longhurst, J. K., Hooyman, A., Albrecht, F., Franzén, E. & Peterson, D. S. Discordance Between Balance Ability and Perception and Its Relation to Falls in Parkinson’s Disease: A Replication Analysis. Neurorehabil. Neural Repair 39, 555–558 (2025).

15. Conklin, S. J. et al. Identifying Predictors of Discordance Between Balance Perception and Ability in Parkinson Disease. J. Neurol. Phys. Ther. https://doi.org/10.1097/NPT.0000000000000556 (2026) doi:10.1097/NPT.0000000000000556.

16. Peterson, D. S. et al. Discordance between actual and perceived balance ability relates to quality of life and global cognition in a clinical sample of Parkinson patients. J. Park. Dis. 16, 529–538 (2026).

17. Albrecht, F. et al. Sedentary behavior modifies the effect of balance rehabilitation on balance discordance in Parkinson’s disease. Npj Park. Dis. 12, 98 (2026).

18. Zhang, B., Zhang, S., Feng, J. & Zhang, S. Age-level bias correction in brain age prediction. NeuroImage Clin. 37, 103319 (2023).

19. ATS Committee on Proficiency Standards for Clinical Pulmonary Function Laboratories. ATS statement: guidelines for the six-minute walk test. Am. J. Respir. Crit. Care Med. 166, 111– 117 (2002).

20. Fulk, G. D., He, Y., Boyne, P. & Dunning, K. Predicting Home and Community Walking Activity Poststroke. Stroke 48, 406–411 (2017).

21. Yang, R. & Pepper, G. A. Is fall self-efficacy an independent predictor of recurrent fall events in older adults? Evidence from a 1-year prospective study. Res. Nurs. Health 43, 602–609 (2020).

22. Pua, Y.-H., Ong, P.-H., Clark, R. A., Matcher, D. B. & Lim, E. C.-W. Falls efficacy, postural balance, and risk for falls in older adults with falls-related emergency department visits: prospective cohort study. BMC Geriatr. 17, 291 (2017).

23. Kato, C., Ida, K. & Harada, A. Influence of high fall-related self-efficacy on falls due to dissociation with ADL among elderly women in nursing homes. Nihon Ronen Igakkai Zasshi Jpn. J. Geriatr. 46, 428–435 (2009).

24. Morone, G., Iosa, M., Pratesi, L. & Paolucci, S. Can overestimation of walking ability increase the risk of falls in people in the subacute stage after stroke on their return home? Gait Posture 39, 965–970 (2014).

25. Sakai, K., Hosoi, Y., Harada, Y., Ikeda, Y. & Tanabe, J. Overestimation associated with walking and balance function in individuals diagnosed with a stroke. Physiother. Theory Pract. 40, 1404–1411 (2024).

26. Takatori, K., Shomoto, K. & Shimada, T. Relationship between Self-Perceived Postural Limits and Falls among Hospitalized Stroke Patients. J. Phys. Ther. Sci. 21, 29–35 (2009).

27. Sakurai, R. et al. Age-related self-overestimation of step-over ability in healthy older adults and its relationship to fall risk. BMC Geriatr. 13, 44 (2013).

28. Byrd, E. M., Jablonski, R. J. & Vance, D. E. Understanding Anosognosia for Hemiplegia After Stroke. Rehabil. Nurs. Off. J. Assoc. Rehabil. Nurses 45, 3–15 (2020).

29. Le, T. T. et al. A Nonlinear Simulation Framework Supports Adjusting for Age When Analyzing BrainAGE. Front. Aging Neurosci. 10, 317 (2018).

30. Butler, E. R. et al. Pitfalls in brain age analyses. Hum. Brain Mapp. 42, 4092–4101 (2021).

31. De Lange, A.-M. G. & Cole, J. H. Commentary: Correction procedures in brain-age prediction. NeuroImage Clin. 26, 102229 (2020).

